# TARGET-HF: Developing a model for detecting incident heart failure among symptomatic patients in general practice using routine health care data

**DOI:** 10.1101/2022.03.17.22270808

**Authors:** Lukas De Clercq, Martijn C. Schut, Patrick Bossuyt, Henk van Weert, M. Louis Handoko, Ralf Harskamp

## Abstract

**Background:** Timely diagnosis of heart failure (HF) is essential to optimize treatment opportunities that improve symptoms, quality of life, and survival. While most patients consult their general practitioner (GP) prior to HF, early stages of HF may be difficult to identify. An integrated clinical support tool may aid in identifying patients at high risk of HF. We therefore constructed a prediction model using routine health care data.

**Methods:** Our study involved a dynamic cohort of patients (≥35 years) who consulted their GP with either dyspnea and/or peripheral edema within the Amsterdam metropolitan area in 2011-2020. **The outcome of interest** was incident HF, verified by an expert panel. We developed a regularized multivariable proportional hazards model (TARGET-HF). The model was evaluated with bootstrapping on an isolated validation set and compared to an existing model developed with hospital insurance data as well as patient age as a sole predictor.

**Results:** Data from 31,905 patients were included (40% male, median age 60) of whom 1,301 (4.1%) were diagnosed with HF over 124,676 person-years of follow-up. Data were allocated to a development (n=25,524) and validation (n=6,381) set. TARGET-HF attained a C-statistic of 0.853 (95%-CI:0.834-0.872) on the validation set, which proved to provide a better discrimination than C=0.822 for age alone (95% CI:0.801-0.842, p<0.001) and C=0.824 for the hospital-based model (95% CI:0.802-0.843, p<0.001).

**Conclusion:** The TARGET-HF model illustrates that routine consultation codes can be used to build a performant model to identify patients at risk for HF at time of GP consultation.

## Background

Heart failure (HF) is a syndrome characterized by the heart’s inability to meet the metabolic needs of the body. The underlying conditions are often multifactorial, and may include comorbidities such as coronary artery disease, hypertension, diabetes mellitus, or valvular disease (1, 2). Overall, around 2% of adults are diagnosed with HF, which increases to >10% in those over the age of 70 (3). Median life expectancy after a diagnosis of HF is five years, and the number of patients with HF is expected to double over the next decades. Timely diagnosis is important to allow initiation of treatments that can improve outcomes, both in terms of mortality as well as quality of life. Leaders in the field of HF strongly propose to prioritize the aim of future research efforts towards early detection and treatment of HF in order to alter the course of disease and limit further deterioration (4, 5).

The key to improve HF detection lies in the community, particularly when focused in a population in which HF is at least moderately prevalent (6). In this regard, a good starting population would therefore be patients who consult their general practitioner (GP) with symptoms that may or may not be related to HF. Two hallmark symptoms for HF are shortness of breath (dyspnea) and peripheral edema, both are unfortunately non-specific in nature, more often than not arising from conditions other than HF (7-9). Guidelines recommend that people with symptoms suggestive of HF presenting to their GP have a natriuretic peptide test as well as an electrocardiogram, with referral for imaging and/or cardiologist review when either is abnormal (1). Still, GPs selectively order these tests based on their perception of the patient’s risk, resulting in selective diagnostic verification and diagnostic delay in patients not deemed at-risk. A report by the British Heart Foundation indicates that nearly eight in ten patients had visited their GP over the previous five years with symptoms associated with heart failure, such as breathlessness or swollen ankles, but had not been diagnosed as such prior to emergency hospital admission (10).

A user-friendly diagnostic support system should be developed to aid the GP to improve risk stratification. Unfortunately, existing HF risk prediction models are not fit for this task as they require a set of actions to be performed, including measurement such as an ECG or laboratory tests, and were not developed for diagnostic support purposes (6, 11). Our primary objective was therefore to construct a model for incident HF based on known predictors, yet only employing preregistered/routine care data available in the patients’ electronic primary care health record. We compare this model with an existing insurance claims based model (2) as well with a model employing patient age as sole predictor.

## Methods

We reported this study in accordance with the Transparent Reporting of a Multivariable Prediction Model for Individual Prognosis or Diagnosis statement (12).

### Data Sourcing

Data were collected in a historical, dynamic cohort of patients registered at any of the general practitioners (GP) affiliated with the *Academic Huisartsennetwerk Amsterdam*, a non-profit organization in charge of region-based acquisition of general practice data for research purposes. The dataset acquired in this network encompasses routine care data from over 600,000 registered patients of 50 practices across sub-municipalities in the cities of Amsterdam and Almere, the Netherlands. Patients at participating practices are able to opt out of this data acquisition process at any point in time. The database is housed on a secure server of the Department of General Practice of the Amsterdam University Medical Center. Researchers can perform analyses on pseudonymized subsets on a separate secure digital environment. Our dataset consists of patients’ demographics, previous consultations, and recurring issues/chronic conditions dubbed “Episodes”. Each consultation’s reason (complaint, symptom, or condition) is coded using the International Classification of Primary Care (ICPC) (13). Due to the nature of primary care, where patients can move to a practice outside the reach of the network, follow-up is not guaranteed across any time period.

### Participants

Our historical cohort has an entry date of 01/01/2011 and an exit date of 31/12/2020, for a total inclusion period of 10 years. It includes all patients registered in the database at the time of extraction (02/06/2021) who complied with the following three criteria: A) the patient has a consultation with an ICPC code for dyspnea (K02, R02) or edema (K07) occurring within the inclusion period at a point where the patient is 35 years or older (the “reference” consultation) with B) at least one consultation thereafter and C) no registered HF diagnoses before the reference consultation.

### Outcome of interest

Potential HF diagnoses found in the patient records were verified manually under close scrutiny of a panel (L.D.C, R.H.) with expertise in general practice, cardiovascular medicine, and medical data processing. A specialized search was initiated for identification of HF diagnoses, where episode records were evaluated for both ICPC codes (K77, K84.03) as well as a series of textual searches of the GP’s notes for terms indicating a HF diagnosis. The regular expressions used for the textual search can be found in Supplement S1. Episodes with a HF code match were deemed a valid incident HF diagnosis unless the accompanying notes indicate otherwise (e.g. expression of doubt, differential diagnoses, …); those with only a textual match were considered invalid unless the context of said match proved otherwise.

### Predictors

We searched the literature for population-based heart failure hazard prediction models to identify variables of interest. Based on the risk factors identified by two systematic reviews, reported by Yang et al. (6) and by Sahle et al. (11), and the availability thereof as ICPC-codes we arrive at a set of 2 demographic values—sex and age—and 14 medical history variables: tobacco use, alcohol abuse, obesity, material deprivation, family history of CVD, hypertension, diabetes mellitus, coronary artery disease, atrial fibrillation, heart murmur, valvular heart disease, stroke, chronic obstructive pulmonary disease, and chronic kidney disease. The full risk factor identification procedure is detailed in Supplement S2.

All conditions prior to each patient’s reference consultation were considered antecedent conditions. The presence thereof was established for the identified risk factors through the means of an ICPC code search, yielding a structured medical history suitable for our regression models. All relevant ICPC codes are detailed in Supplement S3.

### Missing data

The variables regarding medical history represent the registration of conditions rather than the actual presence or diagnosis thereof. In that sense they can—per definition—not be missing. Demographic values are entered for each patient as part of their registration with a GP practice and were found to be complete for all included patients.

### Statistical analysis methods

We calculated incidence of first registered HF diagnoses in our cohort as events per 1,000 person-years, which we report for men and women separately and over different age ranges modelled after those found in Goyal et al. (2). Patients were censored from incidence calculations for the time period beyond their heart failure diagnosis, when present. Incidence rate ratios between sexes are reported across the age groups with their exact Poisson confidence intervals.

A 20% outcome-stratified validation set of our cohort was isolated before the model development process to validate the acquired model. A Cox proportional hazards model was chosen for its ability to handle right-censored data. We fitted our non-parametrical cumulative baseline hazard function using Breslow’s estimator. Taking heed of the concerns Sahle et al. had regarding previous incident HF risk prediction models, we attempted to avoid ill-advised model development practices such as stepwise variable selection or categorization of continuous variables (11).

Similar to Goyal et al. we modeled hazard ratios under three conditions: independently, adjusted for sex and age, and adjusted for all included variables (2). The fully corrected model was subjected to two different forms of regularization and feature selection: using an L_1/2_ penalty (14, 15) and an L_1_ penalty (LASSO) (16). Optimal penalty parameters were selected after a randomized search was applied to a 5-fold cross-validation. A final model candidate for each regularization method was then trained on the entire development set with the acquired penalty values. From these candidates (unregularized, L_1/2_-regularized, L_1_-regularized) the winning model was selected based on calibration statistics where we established the mean (ICI), median (E50), and 90^th^ percentile (E90) of the differences between the calculated survival probabilities and observed proportions on a 1-year horizon. For generating this calibration curve we followed the 3-knot spline procedure as described by Austin et al. (17). All models were constructed and evaluated in Python using the Lifelines (18) and Scikit-survival (19) libraries.

Predictive performance of the chosen model was compared to a baseline of age as a sole predictor as well as the “outpatient” model generated by Goyal et al. based on hospital insurance data. The latter was chosen for its use of routine care data, i.e. not requiring additional measurements or tests, all of which were available as ICPC-codes in our dataset. All models were evaluated using Harrell’s C-statistic (20) on the validation set as a whole as well as stratified by age groups modelled after those used by Goyal et al. Following the principle of Heagerty and Zheng (21) and the weighted method of Uno et al. (22) we calculated area under the cumulative/dynamic receiver-operator curve (AUROC^C,D^) over the first five years of follow-up in one-month increments. Confidence intervals were calculated through bootstrapping (1,000 iterations).

## Results

### Cohort Analysis

A total of 31,905 patients met our cohort criteria, a flowchart of which can be found in Figure S1. Our cohort was 40% male and the median age was 60; further baseline characteristics are summarized in Table 1.

**Table 1:** Baseline characteristics of patients with and without heart failure.

|  | HF<br>(n=1,301) | No HF<br>(n=30,604) | p-value |
| --- | --- | --- | --- |
| Age* (median, 25-75<sup>th</sup>) | 77 (67-84) | 59 (49-71) | <0.001 |
| Years of follow-up* (median, 25-75<sup>th</sup>) | 4.2 (2.2-6.2) | 3.6 (1.5-6.1) | <0.001 |
| Male sex | 601 (46.2%) | 12,047 (39.4%) | <0.001 |
| Tobacco use | 101 (07.8%) | 3,320 (10.9%) | <0.001 |
| Alcohol abuse | 66 (05.1%) | 1,231 (04.0%) | 0.062 |
| Obesity | 141 (10.8%) | 3,950 (12.9%) | 0.028 |
| Material deprivation | 8 (00.6%) | 445 (01.5%) | 0.008 |
| Family history of CVD | 3 (00.2%) | 162 (00.5%) | 0.167 |
| Hypertension | 735 (56.5%) | 10,650 (34.8%) | <0.001 |
| Diabetes mellitus | 433 (33.3%) | 5,170 (16.9%) | <0.001 |
| Coronary artery disease | 404 (31.1%) | 3,507 (11.5%) | <0.001 |
| Atrial fibrillation | 288 (22.1%) | 1,598 (05.2%) | <0.001 |
| Heart murmur | 17 (01.3%) | 261 (00.9%) | 0.092 |
| Valvular heart disease | 167 (12.8%) | 1,025 (03.4%) | <0.001 |
| Stroke | 144 (11.1%) | 1,431 (04.7%) | <0.001 |
| Chronic obstructive pulmonary disease | 256 (19.7%) | 2,806 (09.2%) | <0.001 |
| Chronic kidney disease | 229 (17.6%) | 1,911 (06.2%) | <0.001 |

*Distributional differences are evaluated with Fisher's exact test for proportions and a Mann-Whitney U-test for durations. \*Durations are established relative to the reference consultation, they are reported with median and 25-75th percentiles.*

### Heart Failure Incidence

Inspection of the episodes yielded a total of 4,731 ICPC-code matches and 1,033 additional textual matches, with 698 (8.3%) and 393 (29.5%) of those, respectively, identified as false positive. The textual searches found an additional 10.9% of patients with a HF diagnosis when compared to searching on ICPC-codes alone, as can be seen in Figure S1. After removal of the 2,129 patients with diagnosed heart failure prior to their reference consultation we were left with 1,301 patients (4.08%) diagnosed with HF over 124,676 person-years of follow-up in our cohort.

We observed a HF incidence rate of 10.44 per 1000 person-years (95% CI, 9.88 to 11.02), with 12.96 per 1000 person-years for men (95% CI, 11.95 to 14.04) and 8.94 (95% CI, 8.29 to 9.63) and women. A full overview of rates and rate ratios across sexes and age ranges can be found in supplemental Table S4.

### Predictors and Model Candidates

Our development set for training the models held data from 25,524 persons, of which 1041 (4.1%) had a registered incident HF event in their follow-up period. With a total of 16 variables this puts our models (without feature selection) at over 65 events-per-variable. The resulting hazard coefficients and their respective p-values are reported in supplementary Table S5.

The assessment of calibration for the three fully-adjusted model candidates yielded slight differences, displayed in Figure 1. The L_1_-regularized model displayed overall lower differences between predicted and observed probabilities than its competitors, as can be seen in Table S6. Because of its superior performance—with fewer variables—we opted for the L_1_-regularized model as our final model, designated “TARGET-HF”. Due to the LASSO constraint two variables—tobacco use and family history of CVD—had their coefficients reduced to zero and were effectively eliminated from the model. The calculated baseline hazard is displayed in Figure 2. Hazard ratios for this model, in comparison against those from Goyal et al.’s outpatient model, are shown in Figure 3.

**Figure 1:**
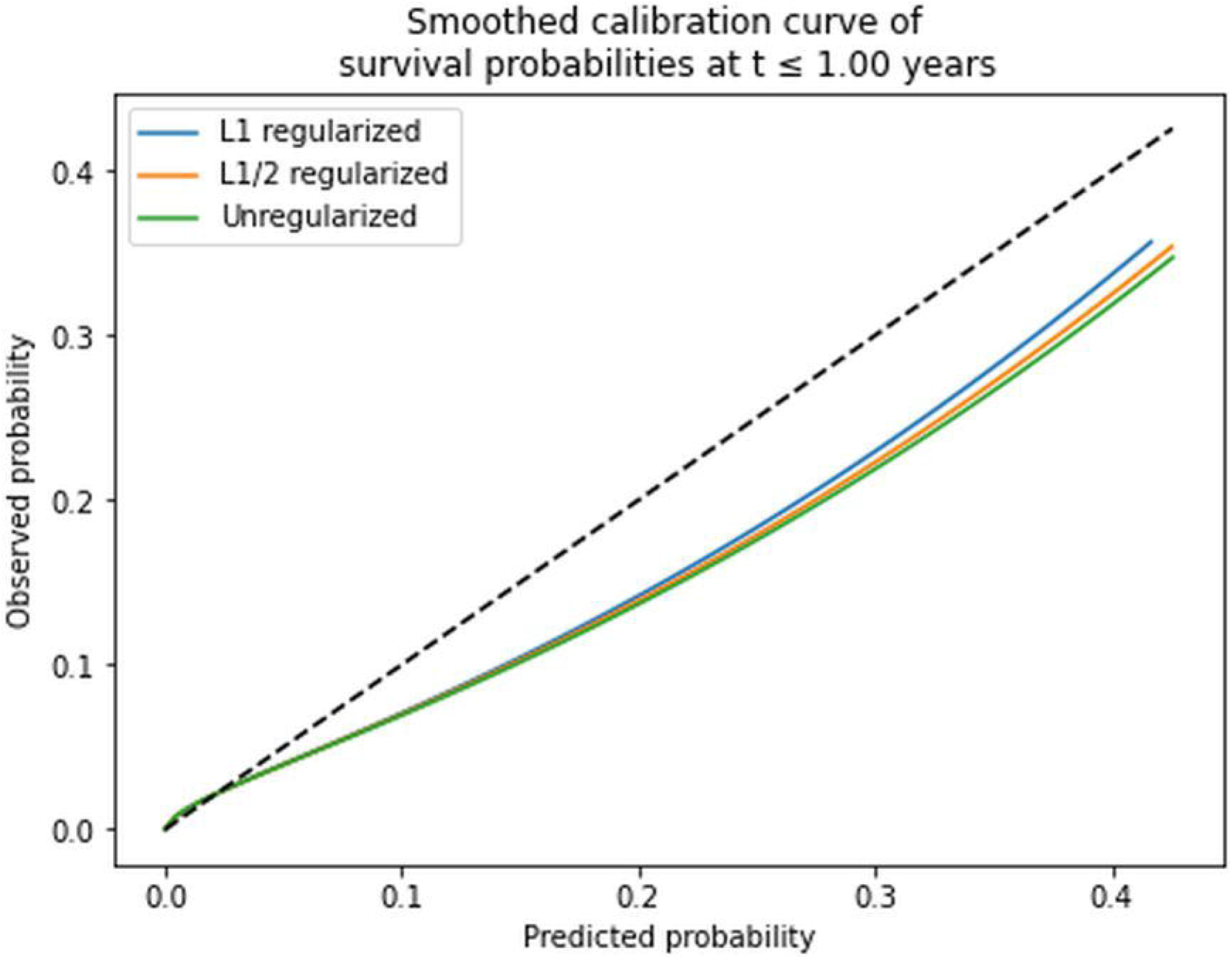
Calibration curves for the three competing models.

**Figure 2:**
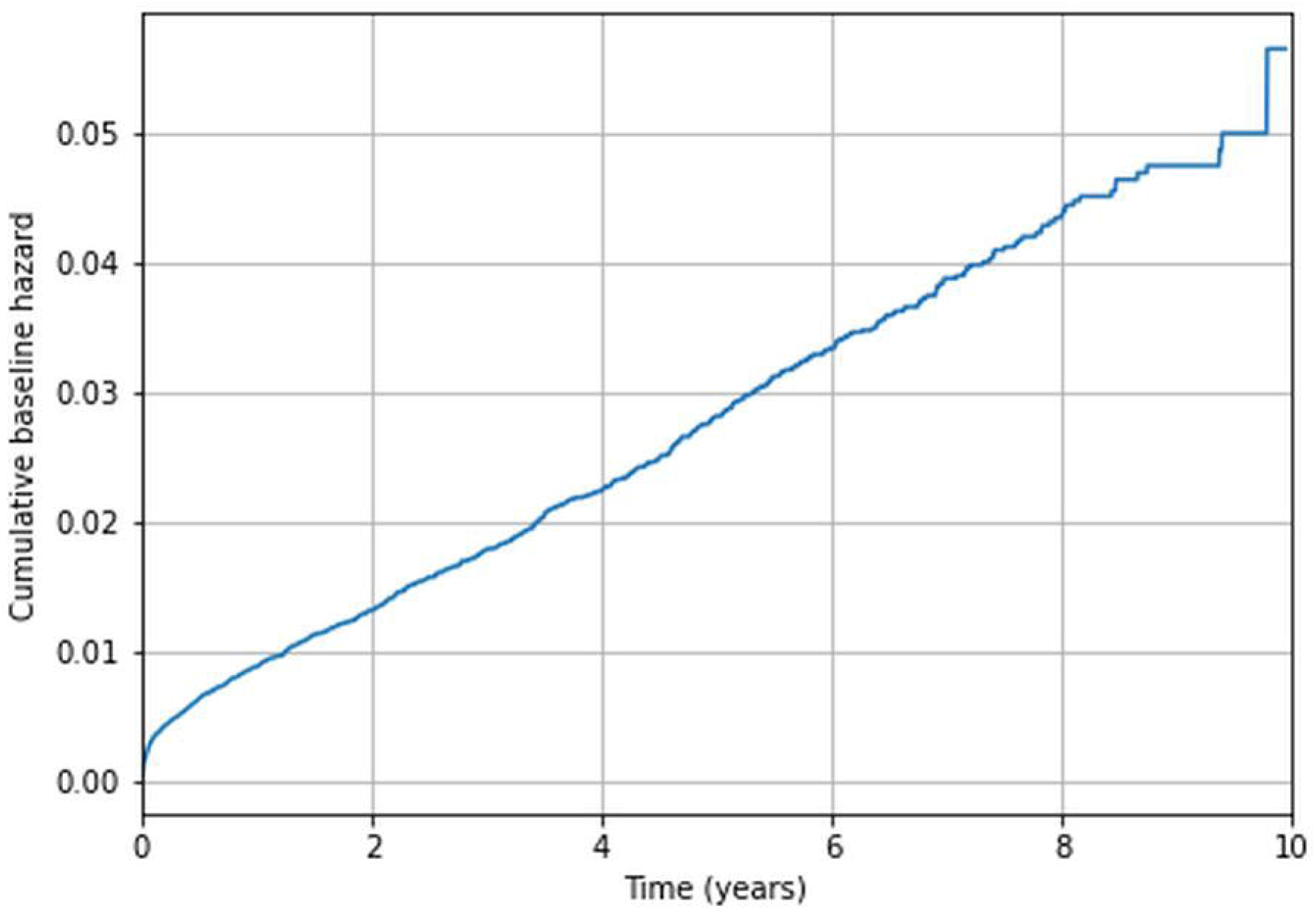
Cumulative baseline hazard fort the L1-penalized proportional hazard model estimated according to Breslow’s method.

**Figure 3:**
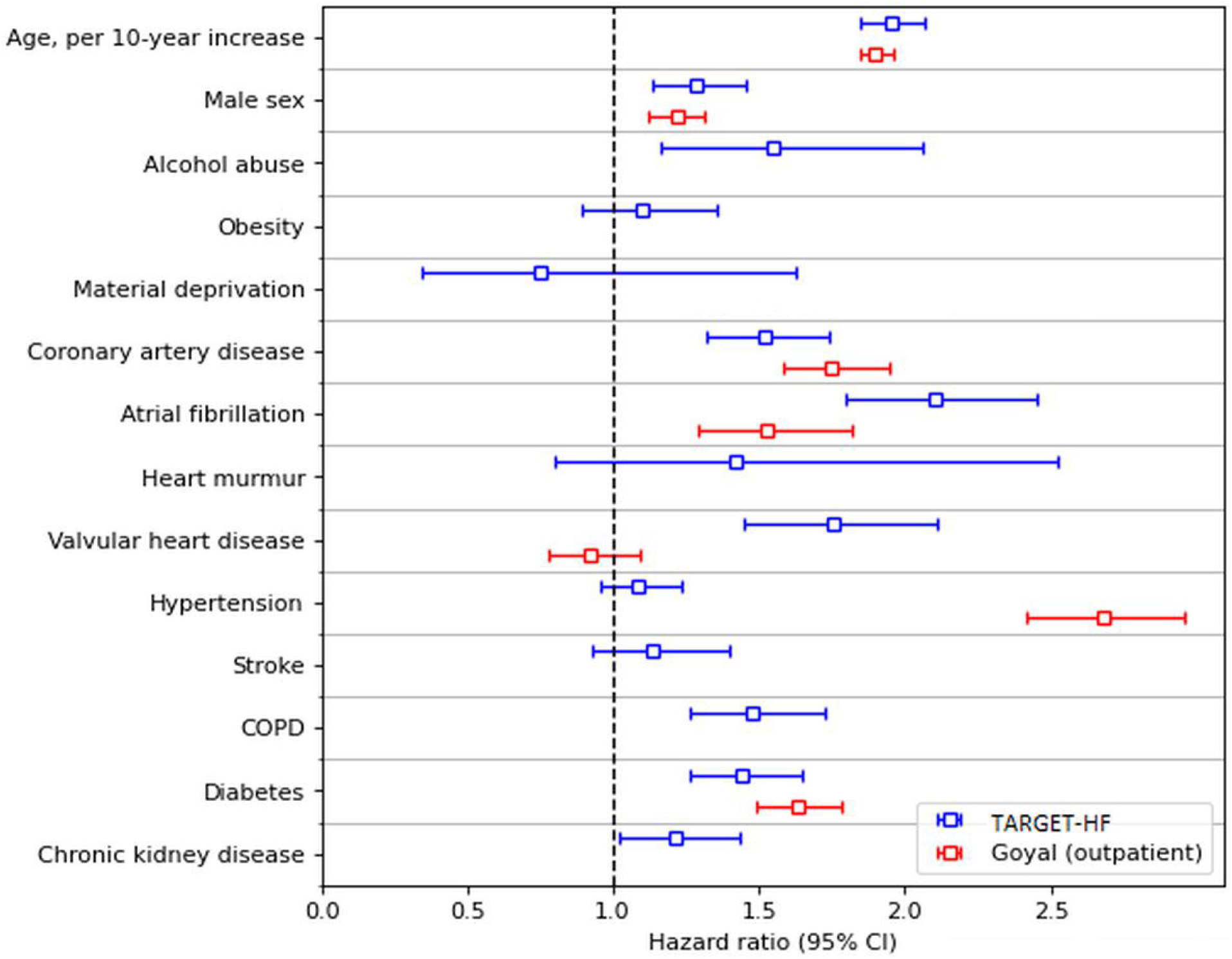
Hazard ratio comparison between our model (TARGET-HF) and Goyal et al.’s outpatient model.

### Predictive Performance

Our validation set held data from 6,381 persons, of which 260 (4.07%) carried an incident HF event during their follow-up period. For this set Harrel’s C-statistic was 0.853 for TARGET-HF (95% CI: 0.834 to 0.872), which proves to outperform C=0.824 for Goyal et al.’s model (95% CI: 0.802 to 0.843, p<0.001) and C=0.822 for the baseline model with age only (95% CI: 0.801 to 0.842, p<0.001). Further C-statistics of the validation set stratified by age can be found in Table 2. Classification performances and their confidence intervals across time in the form of the AUROC^C,D^ can be observed in Table 3 and Figure 4.

**Table 2:** C-statistic of predicting incident heart failure on the validation set as a whole and subdivided by age groups, evaluated for two models and compared to the baseline (age).

| C-statistic | Baseline<br>(age) | TARGET-HF<br>(L<sub>1</sub>-regularized) | Goyal et al.<br>(outpatient) |
| --- | --- | --- | --- |
| All ages | 0.822 (0.801-0.842) | **0.853** (0.834-0.872) | 0.824 (0.803-0.843) |
| 35-54 | 0.721 (0.510-0.857) | 0.831 (0.627-0.943) | **0.833** (0.778-0.888) |
| 55-64 | 0.538 (0.438-0.645) | **0.718** (0.606-0.814) | 0.650 (0.551-0.739) |
| 65-74 | 0.638 (0.572-0.693) | **0.732** (0.662-0.792) | 0.631 (0.568-0.693) |
| 75+ | 0.632 (0.581-0.678) | **0.688** (0.576-0.678) | 0.626 (0.581-0.678) |

**Table 3:**
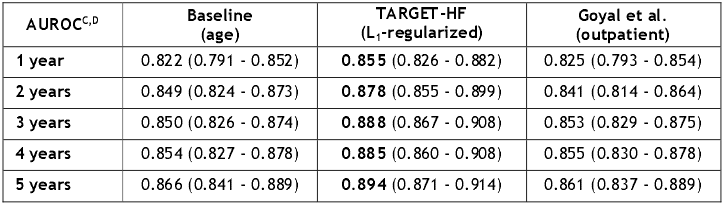
AUROC^C,D^ for three models in predicting incident heart failure in the validation set

| AUROC<sup>C,D</sup> | Baseline<br>(age) | TARGET-HF<br>(L<sub>1</sub>-regularized) | Goyal et al.<br>(outpatient) |
| --- | --- | --- | --- |
| **1 year** | 0.822 (0.791 - 0.852) | **0.855** (0.826 - 0.882) | 0.825 (0.793 - 0.854) |
| **2 years** | 0.849 (0.824 - 0.873) | **0.878** (0.855 - 0.899) | 0.841 (0.814 - 0.864) |
| **3 years** | 0.850 (0.826 - 0.874) | **0.888** (0.867 - 0.908) | 0.853 (0.829 - 0.875) |
| **4 years** | 0.854 (0.827 - 0.878) | **0.885** (0.860 - 0.908) | 0.855 (0.830 - 0.878) |
| **5 years** | 0.866 (0.841 - 0.889) | **0.894** (0.871 - 0.914) | 0.861 (0.837 - 0.889) |

**Figure 4:**
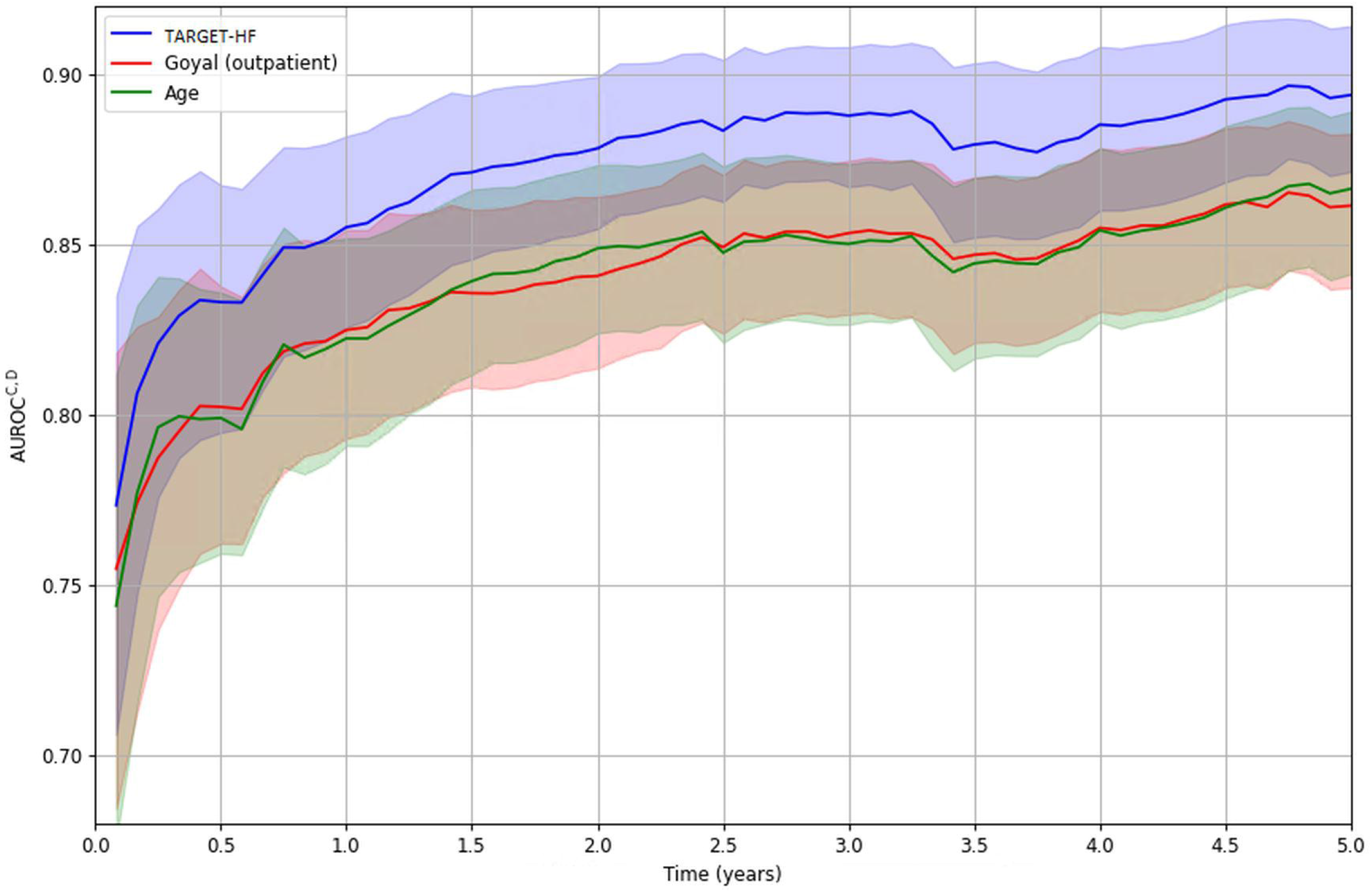
AUROC^C,D^ of TARGET-HF compared to Goyal et al.’s outpatient model and age as a sole predictor in predicting incident heart failure, calculated on a 1-month resolution.

The C-statistic within the development set was 0.812 for TARGET-HF (95% CI: 0.799 to 0.826), outperforming C=0.788 for Goyal et al.’s model (95% CI: 0.775 to 0.803, p<0.001) and C=0.784 for a baseline model based on age alone (95% CI: 0.770 to 0.798, p<0.001).

## Discussion

Timely detection of heart failure starts with accurate identification of patients who should undergo further diagnostic work-up. On this premise, we developed the TARGET-HF model using routine primary care data. Overall, we demonstrated that it is feasible to construct a prediction model that outperforms both age as a sole predictor and an existing community-based prediction model. Moreover, this discriminatory advantage remained consistent when the model was applied to an isolated validation set, notably so in the higher age ranges.

### Strengths and limitations

Our study has a number of strengths. The advantage of using routine care data is that it involves an unselected and therefore representative sample of a population in a metropolitan area. Moreover the data are rich in content due to the availability of structured as well as unstructured elements, such as consultation notes and abstracts of specialist letters, which allowed us to more accurately identify heart failure cases than based on diagnostic codes alone.

Working with routine care data also comes with challenges. First, our methods for identifying heart failure in the follow-up were rigorous but not fool-proof, the large ratio of discarded episodes based on their content gave rise to suspicion towards episodes lacking elaborate descriptions. Few opportunities remain to remediate this situation aside from an external linkage with hospital registrations, though this is hampered by privacy concerns and the lack of a centralized medical record or unique identifier. Second, our predictors are proxies for incidence of the conditions they represent at any point during observation. This means that we did not take into account the time since a code was registered. There may be performance gains in using algorithms capable of modelling this temporal information. A related limitation is the lack of correction for patients’ differing inclinations and/or motives to visit their GP and/or report symptoms or lifestyle complaints. Feasible variables to include to account for this are various derivations of consultation count/frequency prior to the reference consultation.

Selective reporting may also be an issue. Our predictors—the registration of a code at any point in the past—are used as such and may or may not reflect presence or absence of the conditions they represent. In that sense they have to be considered proxies. Especially for lifestyle risk factors there is a concern for underreporting and reporting bias, as they will only be registered when the patient brings these subjects up with their GP or when the GP chooses to register a lifestyle-related code for a consultation for a related complaint. This is exemplified by lifetime tobacco use, which in our cohort is only around 11% for both men and women, whereas a large-scale questionnaire established a far larger proportion in a population sample (23). Nonetheless, these proxies appear to be able to function as predictors for incident heart failure, as shown by our models.

### Prior studies

To our knowledge this study is the first to build a prediction model on HF using routine primary care data. However, community-based models do exist that predict HF. A systematic review of these models found that the strongest associations with incident HF have been observed for age, coronary artery disease, diabetes mellitus, hypertension, and smoking (6). Moreover, atrial fibrillation and valvular heart disease were also strongly associated with HF. When comparing these risk factors with the findings from our analysis we observe similar associations, especially for the demographic values. There were also notable differences, for instance our model’s low weight on hypertension, a factor that typically contributes fairly heavily in other incident heart failure models (6). Another interesting observation is that reported material deprivation yields a coefficient indicating a lower risk compared to the baseline hazard, which contradicts prior findings indicating that material deprivation should be viewed as a risk factor for HF (24). While speculative, we postulate that this may be related to care avoidance and subsequent delayed diagnoses, as well as fewer diagnostic procedures and/or poor registration of incident HF in those with recorded material deprivation versus those without.

#### Clinical implications

HF is a major health problem of increasing prevalence that severely impacts quality of life and shortens lives. (25) It is often not diagnosed early enough to take full advantage of ameliorating medication. (26) In essence, a reliable tool that can help detect high-risk patients for HF in the community would be an important asset to tackle this major health problem.With the model we developed we may provide GPs with an important first building block to improve early detection of HF. The model could be easily integrated in the software of existing primary care electronic health record systems, where it can run as an algorithm in the background to be triggered in a subpopulation of patients presenting with symptoms suggestive for HF. In essence, it could perhaps work in tandem with the recent BEAT-HF campaign, which aims to increase awareness for HF among clinicians in anyone who presents with breathlessness, exhaustion and ankle swelling (5).

#### Future studies

The prototype that we have developed appears promising, but further validation is warranted prior to implementation. Moreover, interventional studies would then be required to evaluate whether integrating this algorithm in GP practice, indeed results in improved HF detection, initiation of adequate treatment and ultimately improved clinical outcomes. However, prior to these steps, we would propose to evaluate whether additional techniques, such as natural language processing and other machine learning techniques, would even further improve prediction of HF, and may also help to further improve accurate diagnostic coding. As our study showed, textual queries provide us with an additional 10% of HF diagnoses not caught by ICPC codes, a non-negligible number for such a serious heart condition. In other words, there is still work to be done and we believe that there may be predictive potential in the unstructured data of GP patient files that has thus far been untapped, which requires a different approach to be used to its full potential.

### Conclusion

The TARGET-HF model illustrates that consultation codes found in routine primary care data can be used to build a performant model to identify patients at risk for heart failure at time of GP consultation. Moreover, we found that the model outperformed both age as a sole predictor as well as an existing community-based prediction model based on hospital insurance data.

## Supporting information

TRIPOD checklist

## Data Availability

All data produced in the present work are contained in the manuscript

## Conflicts of interest

None

## Financial support

This work was supported by the department of general practice of the Amsterdam UMC – AMC location, as well as by a grant of the Amsterdam Public Health Research Institute.

## Notes

### Competing Interest Statement

The authors have declared no competing interest.

### Author Declarations

Ethics Amsterdam UMC - waived

