## Supplemental data for "TARGET-HF: Developing a model for detecting incident heart failure among symptomatic patients in general practice using routine health care data"

### Supplementary materials

#### Supplement S1: Regular expressions used for HF episode search

- hart *falen
- cardio *m[iy]opath?ie
- dec(\.?|omp\w*\.?) *cordis

Either of the queries above qualified for a match unless they were directly followed by a question mark (indicating uncertainty) or preceded by either of the following terms indicating negation, doubt, or an expression of fear for a diagnosis:

- geen
- niet
- zonder
- mogelijk
- verdenking
- cave
- dd
- d.d.
- angst voor
- schrik voor

#### Supplement S2: Risk factor identification procedure

We found two systematic reviews by Yang et al. [4] and Sahle et al. [9] that evaluated a series of articles, respectively 15 and 19 (of which 5 overlap), reporting risk factors for incident heart failure. One article, present in both reviews, employs a logistic regression rather than survival analysis model [12] and variables only present there were therefore not included in our consideration. Whereas Yang et al. set an unselected population as an inclusion criterion in their review, Sahle et al. have no such requirement.

Although there are notable differences between the risk factors identified by both reviews, 11 out of 15 medical history variables are shared between the two. For the demographic variables we established whether they are available as structured data in the patients’ records. Note that neither race nor education level are consistently registered in Dutch primary care and when present in a patient record it is not in a structured fashion. When a variable is either present in the patient record or has a corresponding ICPC code it qualified for our model input. Due to the presence of the superclass *coronary artery disease* two of its subclasses, *angina pectoris* and *myocardial infarctions*, were not considered as regression models can produce unstable results with highly correlated predictors. This procedure yielded a total of 2 demographic risk factors and 14 medical history variables with corresponding ICPC codes. All risk factors identified by these reviews are summarized in supplemental Table S3, where the presented risk factors are accompanied by their respective ICPC codes (when available) as identified by the authors.

#### Supplemental Table S3: Summary of identified risk factors

| **Variable** | **Abbreviation** | **Yang et al.** | **Sahle et al.** | **ICPC code** | **Patient record** |
| --- | --- | --- | --- | --- | --- |
| **Demographics** | | | | | |
| Age |  | X | X |  | X |
| Sex |  | X | X |  | X |
| Race |  | X | X |  |  |
| Education level |  | X |  |  |  |
| **Lifestyle** | | | | | |
| Alcohol abuse |  | X |  | P15, P16 |  |
| Tobacco use |  | X | X | P17 |  |
| Obesity |  | X | X | T82, T83 |  |
| Material deprivation |  |  | X | Z01 |  |
| Physically inactive |  | X |  |  |  |
| **Medical history** | | | | | |
| Family history of cardiac disease |  | X |  | A29.01 |  |
| Coronary artery disease |  | X | X | K47, K75, K76 K76.02 |  |
| Atrial fibrillation | AF | X | X | K78 |  |
| Heart murmur |  |  | X | K81 |  |
| Valvular heart disease | VHD | X | X | K83 |  |
| Hypertension | HTN | X | X | K86, K87 |  |
| Stroke |  | X | X | K90 |  |
| Chronic obstructive pulmonary disease | COPD | X | X | R91, R95 |  |
| Diabetes mellitus | DM | X | X | T90 |  |
| Chronic kidney disease | CKD | X | X | U99.01 |  |
| **Observations/measurements** |  |  |  |  |  |
| Abnormal ECG* |  | X | X |  |  |
| Left ventricular hypertrophy | LVH | X | X |  |  |
| Body mass index | BMI |  | X |  |  |
| Microalbuminuria |  |  | X |  |  |
| Dyslipidemia |  | X |  |  |  |
| Systolic blood pressure | SBP |  | X |  |  |
| Diastolic blood pressure | DBP |  | X |  |  |
| Resting heart rate | RHR | X | X |  |  |
| Left ventricular ejection fraction |  | X |  |  |  |
| **Medication use** |  |  |  |  |  |
| Antihypertensive |  |  | X |  |  |
| Pioglitazone |  |  | X |  |  |
| **Biomarkers** |  |  |  |  |  |
| Low-density lipoprotein | LDL |  | X |  |  |
| High-density lipoprotein | HDL |  | X |  |  |
| Total cholesterol | TC |  | X |  |  |
| TC/HDL ratio |  |  | X |  |  |
| Fasting glucose | FBG | X | X |  |  |
| C-reactive protein | CRP | X | X |  |  |
| Hemoglobin | Hb |  | X |  |  |
| Hemoglobin A1c | HbA1c |  | X |  |  |
| Albumin | Alb | X | X |  |  |
| Creatinine | Cr | X | X |  |  |
| Albumin/creatinine ratio | ACR |  | X |  |  |
| Brain natriuretic peptide | BNP |  | X |  |  |
| N-terminal pro-brain natriuretic peptide | NT-proBNP | X | X |  |  |
| Mid-regional pro-atrial natriuretic peptide | MR-proANP |  | X |  |  |
| Interleukin 6 | IL-6 |  | X |  |  |
| Tumor necrosis factor alpha | TNF-α |  | X |  |  |
| Soluble interleukin 1 receptor-like 1 | sST2 |  | X |  |  |
| Troponin(T, C, I) |  |  | X |  |  |
| Growth/Differentiation Factor-15 | GDF15 |  | X |  |  |
| Cystatin C | CysC |  | X |  |  |
| Galectin-3 | Gal-3 |  | X |  |  |
| Procalcitonin | PCT |  | X |  |  |

*Risk factors for incident heart failure in the population from Yang et al. and Sahle et al. *Different models employed different (combinations of) ECG abnormalities, none of which have a corresponding ICPC code.*

#### Supplemental Table S4: Heart Failure Incidence

|  | | **35-54 years** | **55-64 years** | **65-74 years** | **≥75 years** | **Total** |
| --- | --- | --- | --- | --- | --- | --- |
| Incident HF cases | Male | 43 | 74 | 131 | 353 | 601 |
|  | Female | 30 | 84 | 125 | 461 | 700 |
|  | Total | 73 | 158 | 256 | 814 | 1301 |
| Person-years of follow-up | Male | 14,234 | 13,241 | 10,759 | 8,134 | 46,367 |
|  | Female | 29,135 | 19,155 | 14,721 | 15,298 | 78,309 |
|  | Total | 43,368 | 32,396 | 25,480 | 23,432 | 124,676 |
| Incidence rate per 1000  person-years | Male | 3.02  (2.19-4.07) | 5.59  (4.39-7.02) | 12.18  (10.18-14.45) | 43.40  (38.99-48.17) | 12.96  (11.95-14.04) |
|  | Female | 1.03  (0.70-1.47) | 4.39  (3.50-5.43) | 8.49  (7.07-10.12) | 30.13  (27.45-33.02) | 8.94  (8.29-9.63) |
|  | Total | 1.68  (1.32-2.12) | 4.88  (4.15-5.70) | 10.07  (8.87-11.38) | 34.74  (32.39-37.21) | 10.44  (9.88-11.02) |
| Male/female  incidence rate ratio | | 2.94  (1.80-4.84) | 1.27  (0.92-1.76) | 1.43  (1.11-1.85 | 1.44  (1.25-1.66) | 1.45  (1.30-1.62) |
| p-value | | <0.0001 | 0.1299 | 0.0040 | <0.0001 | <0.0001 |

*Heart failure incidence, rates and ratios in a cohort of symptomatic patients between 2011 and 2020, subdivided in age ranges and sexes.*

#### Supplemental Table S5: Cox proportional hazards coefficients for incident heart failure.

| **Covariate** | **Unadjusted** | | **Adjusted for sex and age** | | **Adjusted for all covariates** | | **L_1/2_-regularized** | | **L_1_-regularized** | |
| --- | --- | --- | --- | --- | --- | --- | --- | --- | --- | --- |
| Age | 2.23  (2.12 – 2.34) | <0.0001 | - | - | 1.97  (1.87 – 2.09) | <0.0001 | 1.97  (1.87 – 2.09) | <0.0001 | 1.96  (1.85 – 2.07) | <0.0001 |
| Male | 1.45  (1.28 – 1.64) | <0.0001 | - | - | 1.32  (1.16 – 1.49) | <0.0001 | 1.30  (1.15 – 1.47) | <0.0001 | 1.28  (1.13 – 1.46) | <0.0001 |
| Alcohol abuse | 1.44  (1.10 – 1.90) | 0.0087 | 1.72  (1.30 – 2.27) | 0.0001 | 1.64  (1.24 – 2.17) | 0.0006 | 1.61  (1.22 – 2.13) | 0.0009 | 1.55  (1.17 – 2.06) | 0.0025 |
| Tobacco use | 0.73  (0.58 – 0.91) | 0.0061 | 1.10  (0.88 – 1.38) | 0.4076 | 0.97  (0.77 – 1.22) | 0.7709 | - | - | - | - |
| Obesity | 0.74  (0.60 – 0.90) | 0.0027 | 1.27  (1.03 – 1.55) | 0.0242 | 1.17  (0.95 – 1.43) | 0.1461 | 1.13  (0.92 – 1.40) | 0.2386 | 1.10  (0.89 – 1.36) | 0.3686 |
| Material deprivation | 0.35  (0.14 – 0.84) | 0.0189 | 0.67  (0.28 – 1.61) | 0.3678 | 0.58  (0.24 – 1.40) | 0.2246 | 0.63  (0.27 – 1.46) | 0.2860 | 0.75  (0.35 – 1.63) | 0.4660 |
| CVD in family | 0.64  (0.21 – 1.99) | 0.4439 | 1.34  (0.43 – 4.17) | 0.6127 | 1.46  (0.47 – 4.54) | 0.5141 | 1.26  (0.37 – 4.24 | 0.7132 | - | - |
| CAD | 3.38  (2.97 – 3.86) | <0.0001 | 1.70  (1.48 – 1.95) | <0.0001 | 1.53  (1.33 – 1.75) | <0.0001 | 1.53  (1.33 – 1.75) | <0.0001 | 1.52  (1.32 – 1.74) | <0.0001 |
| AF | 5.48  (4.73 – 6.35) | <0.0001 | 2.32  (1.99 – 2.70) | <0.0001 | 2.11  (1.80 – 2.46) | <0.0001 | 2.11  (1.81 – 2.46) | <0.0001 | 2.10  (1.80 – 2.45) | <0.0001 |
| Murmur | 1.46  (0.84 – 2.52) | 0.1782 | 1.51  (0.87 – 2.61) | 0.1406 | 1.57  (0.90 – 2.71) | 0.1098 | 1.52  (0.87 – 2.65) | 0.1409 | 1.42  (0.80 – 2.52) | 0.2330 |
| VHD | 4.37  (3.65 – 5.23) | <0.0001 | 2.08  (1.73 – 2.50) | <0.0001 | 1.77  (1.47 – 2.13) | <0.0001 | 1.77  (1.47 – 2.13) | <0.0001 | 1.75  (1.45 – 2.11) | <0.0001 |
| HTN | 2.38  (2.11 – 2.69) | <0.0001 | 1.25  (1.10 – 1.42) | 0.0005 | 1.10  (0.97 – 1.25) | 0.1302 | 1.09  (0.96 – 1.24) | 0.1961 | 1.09  (0.96 – 1.24) | 0.1941 |
| Stroke | 2.67  (2.18 – 3.26) | <0.0001 | 1.32  (1.08 – 1.61) | 0.0075 | 1.16  (0.95 – 1.42) | 0.1511 | 1.14  (0.93 – 1.40) | 0.1950 | 1.14  (0.93 – 1.40) | 0.2224 |
| COPD | 2.33  (2.00 – 2.72) | <0.0001 | 1.54  (1.32 – 1.80) | <0.0001 | 1.51  (1.29 – 1.77) | <0.0001 | 1.49  (1.28 – 1.75) | <0.0001 | 1.47  (1.26 – 1.73) | <0.0001 |
| DM | 2.44  (2.14 – 2.77) | <0.0001 | 1.57  (1.38 – 1.79) | <0.0001 | 1.46  (1.28 – 1.67) | <0.0001 | 1.46  (1.28 – 1.66) | <0.0001 | 1.44  (1.27 – 1.65) | <0.0001 |
| CKD | 3.70  (3.15 – 4.34) | <0.0001 | 1.41  (1.20 – 1.67) | <0.0001 | 1.22  (1.03 – 1.44) | 0.0208 | 1.21  (1.03 – 1.44) | 0.0243 | 1.21  (1.02 – 1.43) | 0.0261 |

**Supplemental Table S6***: The mean (ICI), median (E50), and 90^th^ percentile (E90) of the differences between the calculated survival probabilities and the observed frequencies of three covariate-adjusted Cox proportional hazards models, calculated on the development set on a 1-year horizon. Lower is better for all statistics.*

| **Calibration statistic (%)** | **Unregularized** | **L_1/2_-regularized** | **L_1_-regularized** |
| --- | --- | --- | --- |
| **ICI** | 0.53 | 0.51 | **0.50** |
| **E50** | 0.32 | 0.31 | **0.30** |
| **E90** | 1.01 | 1.00 | **0.99** |

*Figure S1: Cohort inclusion criteria and heart failure episode verification results. t_HF_ signifies the time of the identified HF diagnosis registration, t_0_ symbolizes the reference consultation.*

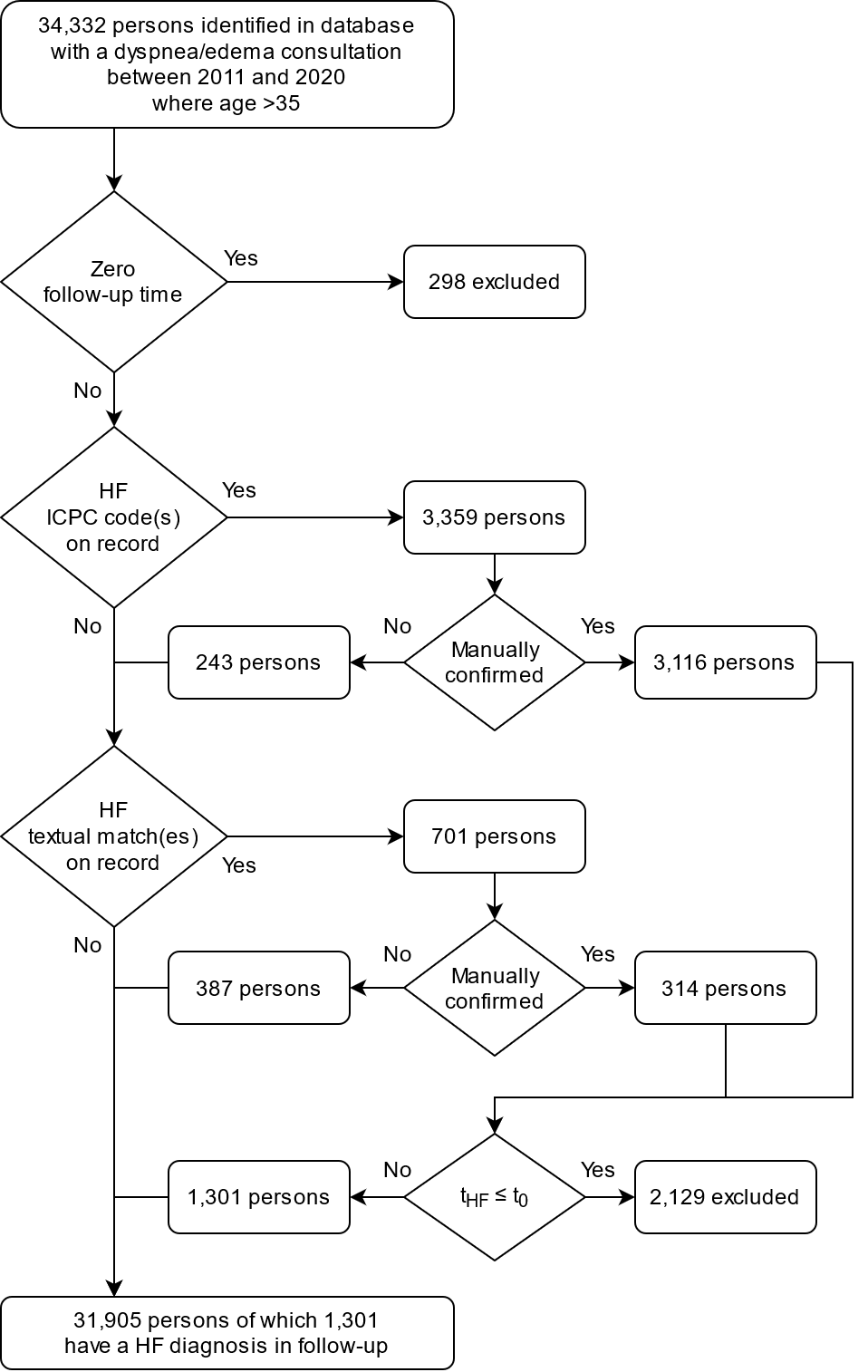
